# Successful incorporation of a genetic risk prediction research platform into routine newborn screening

**DOI:** 10.1101/2021.02.26.21252305

**Authors:** Owen M Bendor-Samuel, Tabitha Wishlade, Louise Willis, Parvinder Aley, Edward Choi, Rachel Craik, Yama Mujadidi, Ginny Mounce, Fenella Roseman, Arancha De La Horra Gozalo, James Bland, Nazia Taj, Ian Smith, Anette-Gabriele Ziegler, Ezio Bonifacio, Christiane Winkler, Florian Haupt, John A. Todd, Laurent Servais, Matthew D Snape, Manu Vatish, the GPPAD Study Group

**Author notes:** **Correspondence:** M Vatish, Nuffield Dept of Women’s & Reproductive Health, John Radcliffe Hospital, Oxford, OX3 9DU, United Kingdom. Joint first authors (Owen M Bendor-Samuel, Tabitha Wishlade). joint senior authors (Matthew D Snape, Manu Vatish).

## Abstract

An increasing number of diseases can be offered treatments that are transformative if administered in a timely manner. However, many of these diseases are currently not included in the newborn screening programs because they lack sensitive and specific metabolic biomarkers, and detection of children at increased risk relies on genetic methods. Type 1 diabetes (T1D) constitutes a potential example of such disease.

Between April 2018 and November 2020, over 15500 babies were enrolled into ‘INGR1D’ (<u>In</u>vestigating <u>G</u>enetic <u>R</u>isk for T<u>1D</u>), a research study to identify newborns with an increased genetic risk of T1D. This project, performed as part of a T1D primary prevention study (the Primary Oral Insulin Trial, POInT), has helped to pioneer the integration of genetic screening into the NHS Newborn Blood Spot Screening Programme (NBSSP) for consenting mothers, without affecting the screening pathway. The use of prospective consent to perform personalised genetic testing on samples obtained through the routine NBSSP represents a novel mechanism for clinical genetic research in the UK and provides a model for further population based genetic studies in the newborn.

This project builds on the UK’s role as a world leader in genomic medicine, e.g. through its inception and completion of the 100 000 Genomes Project, and its subsequent ambition to map 5 million further genomes over the next 5 years.

Our aim is therefore to describe the methodology used by INGR1D as a way to demonstrate how a successful research and clinical trial tool can be integrated into a national screening programme, with the potential for the tool to be developed to incorporate multiple diseases with genetic markers without altering the screening pathway.

## INTRODUCTION

The UK has conducted its newborn screening programme for more than half a century and has been a successful example of mass population based metabolic screening designed to detect diseases with severe morbidity or mortality that are managed more effectively with earlier detection.[1] In that time we have also gained more knowledge about genetic predisposition and genetic markers for disease e.g. Type 1 Diabetes (T1D), Severe Combined Immunodeficiency (SCID) and Spinal Muscular Atrophy (SMA). More recently, therapeutic options for such diseases (either in a research context or as approved therapies) have become available. There is therefore the potential to use a Newborn Blood Spot Screening Programme (NBSSP) to screen for genetic disorders as well as screening for metabolic disease. With this in mind, we describe a successful methodology which allows antenatal consent to be gained for entry of newborns onto a T1D genetic screening study called INGR1D (<u>IN</u>vestigating the <u>G</u>enetic <u>R</u>isk of Type <u>1 D</u>iabetes). Furthermore, this paper explores how it could be adapted for research and evaluation of potential blood spot screening programmes for other important genetic diseases (e.g. SMA).

### SCREENING IN THE UK

Screening newborn infants for metabolic diseases in the United Kingdom was first established in the late 1950s. Initially, the screening programmes were organised locally and utilised a urine test for the early detection of phenylketonuria (PKU). Over the following decade, PKU screening became a national recommendation and the **Newborn blood spot screening card (NBSSC)**, originally known as a Guthrie card, replaced the inferior urine-testing. The NHS NBSSP now screens for nine conditions (Table 1) as selected by the National Screening Committee (NSC), based on a ‘Format’ of 19 criteria.[1]

**Table 1.** List of the 9 conditions in the newborn screening

| Conditions included in newborn screening |  |  |
| --- | --- | --- |
| Medium-chain acyl-CoA dehydrogenase deficiency | Phenylketonuria | Sickle cell and Thalassaemia |
| Maple syrup urine disease | Isovaleric acidemia | Cystic fibrosis |
| Congenital hypothyroidism | Glutaric aciduria type 1 | Homocystinuria |

To undertake this screening, newborn infants have four bloodspots collected at day 5 of life onto a NBSSC. The cards are processed and analysed in 16 central laboratories across the UK where four 3.2mm diameter spots are punched for the conditions currently being screened. If four good quality spots are collected, each card has the potential to provide approximately 16 blood spots. This provides redundancy if cards need to be re-analysed for any patient with positive, borderline or inconclusive results, without needing to re-bleed the infant. It also ensures that the blood spots used for quantitative assays can be punched from an area where the blood is evenly distributed.

This additional redundancy provides the potential for other screening tests to be added, including for research purposes. With an average national coverage of 96.5%, the NBSSP in the UK is widely acceptable to families [2] and provides an ideal platform for recruitment into research studies. Despite its vast potential, as far as the authors are aware, this has never been utilised prospectively on a large scale.

### TYPE 1 DIABETES and INGR1D

T1D is an autoimmune condition that leads to significant mortality and morbidity as a consequence of insulin deficiency and ensuing hyperglycaemia. In 2017 there were 40 300 children and adolescents (<20 years) with T1D in the UK, representing the 7^th^ highest prevalence of T1D globally. The UK also has the world’s 5^th^ highest number of new cases per year of T1D in those younger than 15 years of age, equating to 3300 new cases per year.[3] Moreover, the incidence of T1D has been increasing by 3% year-on-year.[4-7] Peak age of diagnosis is 10-14 years of age [8] and treatment of T1D requires life-long insulin replacement therapy. However, due to the difficulties with maintaining normoglycaemia, life expectancy in 20-year-old diabetics is reduced by 11 and 13 years in men and women respectively.[9]

Beta cells in the islets of Langerhans, responsible for insulin production, are destroyed through an immune-mediated process that can be identified by circulating islet cell autoantibodies. Through a number of T1D observational cohort studies it has become apparent that the break in immune self-tolerance, marked by the presence of islet autoantibodies (IA), occurs as early as 3-6 months of age and peaks at the age of 2 years. In addition, the presence of two or more IA is predictive of T1D, with 80% of individuals developing symptoms over the following 10 years. Individuals with multiple IA can therefore be thought of having an early stage of T1D known as asymptomatic or pre-diabetes.[10-14]

Achieving self-tolerance is facilitated by T-cell exposure of self-antigens in the thymus or secondary lymphoid tissues (such as lymph nodes, gut or spleen), leading to induction of regulatory T cells and deletion of autoreactive effector T cells. The risk of T1D is known to be influenced by polymorphisms in the *INSULIN (INS)* gene that affect insulin expression in the thymus and hence disturb the self-tolerance pathway.[15, 16] This therefore raises the question as to whether such a process could be influenced by inducing self-tolerance through regular oral mucosal exposure of insulin in infancy when immune mechanisms driving tolerance are fully active. In support of this hypothesis, the LEAP trial successfully demonstrated that early and repeated exposure to peanuts can induce tolerance and lead to a sevenfold reduction in the risk of peanut allergy.[17] The Global Platform for the Prevention of Autoimmune Diabetes (GPPAD)[18] is now undertaking a primary prevention trial, called Primary Oral Insulin Trial (POInT),[19] aiming to emulate the success of the LEAP study with early exposure to oral insulin prior to IA seroconversion.

### GENETIC RISK SCORE

Conducting primary prevention clinical trials in T1D have historically been difficult due to the inability to identify an at-risk population large enough to be approached for recruitment. Having a first-degree relative increases the risk of T1D to one-in-twenty. However, 85% of newly diagnosed diabetics do not have a family history of the disease.[20] Solely targeting T1D first-degree relatives (FDR) would therefore miss a large proportion of prospective cases and would require a large geographical footprint to yield an adequate sample size. This problem was resolved by creating a genetic risk score (GRS) based on 47 single nucleotide polymorphisms (SNP) that enables stratifying T1D risk. The scoring system was generated from the T1D Genetic Consortium (T1DGC) and Wellcome Trust Case Control Consortium (WTCCC) and was analysed to identify HLA class II genotypes and 40 non-HLA SNPs associated with T1D risk.[21] Individuals can now be identified as having a 10% risk of developing asymptomatic T1D by 6 years of age by using HLA typing in those with a T1D FDR, or the GRS in conjunction with HLA type in those without a T1D FDR.[18, 22-27]

### SCREENING FOR A RANDOMISED CONTROL TRIAL

Having developed an algorithm to screen for an increased risk of T1D and identified an antigen to induce self-tolerance, a randomised control trial could feasibly be designed. Following this, a safe and immunogenic dose of insulin was identified with the potential to provide a tolerogenic response.[28] GPPAD proceeded to devise two studies, a screening study (known as INGR1D in the UK) that would identify an at-risk population of T1D and a randomised control trial, known as POInT,[19] that would recruit from the aforementioned population. Between October 2017 and November 2020 233,164 newborns were screened across five European countries (Germany, UK, Sweden, Belgium and Poland) and 887 enrolled into the POINT clinical trial, with recruitment expected to cease in March 2021.[21]

### INGR1D Study Design and Initiation

Recruitment to INGR1D began in April 2018 and was completed in November 2020. Approvals for this study were obtained from the National Screening Programme Research Advisory Committee, the Hampshire A Research Ethics Committee and the NHS Research and Development committees of the relevant NHS trusts. There were four INGR1D screening trusts in the UK:

- Oxford University Hospital (OUH) NHS Foundation Trust
- Buckinghamshire Healthcare NHS Trust
- Royal Berkshire NHS Foundation Trust
- Milton Keynes University Hospital NHS Foundation Trust

The recruiting hospitals within these trusts represented the busiest delivery units in the Thames Valley and crucially, used the same NHS newborn blood spot screening laboratory (NBSSL) in Oxford. The majority of participants were recruited by research midwives in antenatal clinics from 18 weeks’ gestation onwards, providing verbal and written information on INGR1D and POInT. If they were willing to participate, consent was taken electronically on a tablet to allow for a) completion of a questionnaire and b) prospective consent to use surplus neonatal screening blood for genetic testing. Additionally, in order to allow enrolment of families from outside the Thames Valley area, or those whose infants had already had their neonatal screening test performed before their parents became aware of the study, recruitment of babies up to 3 months of age was allowed if parents were willing to travel to the Oxford study centre. For these two groups of participants, a blood spot was taken on an additional NBSSC which was clearly labelled as a “GPPAD only” sample. This pathway therefore did not interfere with the child’s routine newborn screening which was undertaken at their regional screening laboratory.

With the advent of the coronavirus pandemic, face-to-face recruitment was temporarily halted at OUH in favour of receiving consent remotely. To facilitate this, a master list of women at the trust who were around 36 weeks pregnant was sent to the recruiting team each week. The team then checked each woman’s electronic hospital record to confirm eligibility for the study. If eligible, a template email or text about the study was sent to women using the contact details on their electronic record. If after 48 hours there was no response, women were telephoned a maximum of twice by the study team. Once contact was established, women who wished to participate could either complete and return an electronic version of the consent form or give verbal consent to recruiting staff over the phone. If there was no response to the phone calls women were presumed to have declined participation in the study and no further contact was sought.

In order to incorporate the T1D genetic screening into the screening process, the Laboratory Information System, OMNI-Lab. (Integrated Software Solutions Ltd), used by the Oxford NBSSL, was modified. The modification facilitated NBSSCs from consented babies to be linked to the study consent list via the mother’s NHS number and to generate a request on the system for a GPPAD test. For babies participating in INGR1D this:

- Alerted laboratory staff to punch an additional 3.2 mm sample which was sent to LGC Biosearch Technologies for genotyping.
- Sent a sample receipt notification to the INGR1D database together with the Child’s date of birth and gender.
- Sent the plate identifier and well position of the spot to LGC.

Results were forwarded to Helmholtz Zentrum München, the coordinating centre (CC) in Munich, which integrated the genotyping data, information from the screening laboratory and maternal questionnaire to generate a genetic risk score (for the detailed process please refer to Figure 1). Mothers were informed of positive results within 16 weeks from the analysis of the sample and subsequently offered a face-to-face appointment to explain the implications of the result and informed about POInT. Mothers of babies with negative results were not specifically advised of the result but were told at the time of consent that a negative result could be inferred if the study team did not contact them by 16 weeks. If parents remained anxious about the result, they could also contact the study team directly. Parents could withdraw their consent at any time.

**Figure 1.**
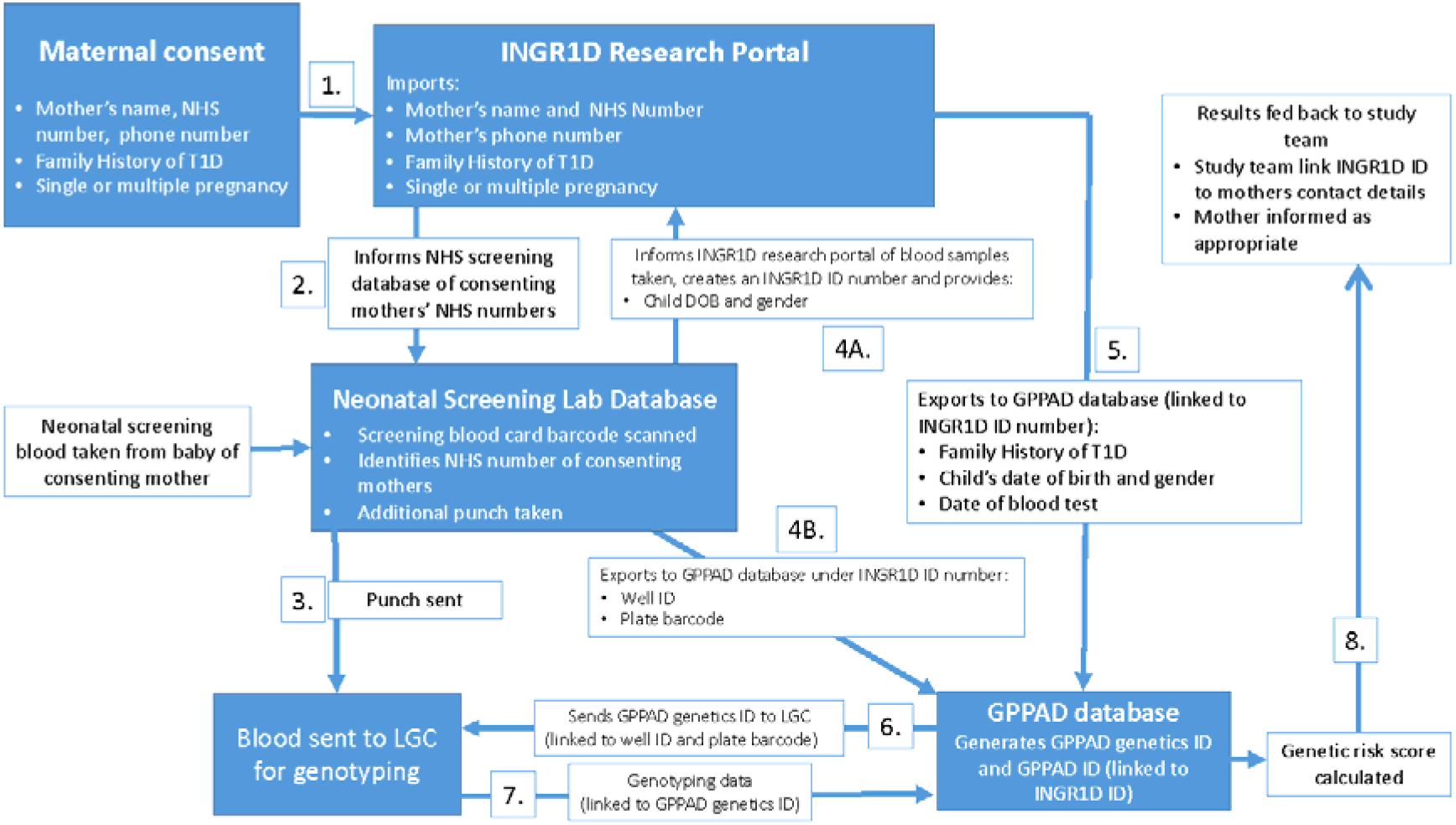
1. The questionnaire asks for the mother’s name, NHS number, phone number, e-mail, family history of T1D, and if they have a single or multiple pregnancy. This information is stored on the **INGR1D research portal**. 2. The **research portal** collates the consented patients and informs the **neonatal screening laboratory** of the consenting mothers’ NHS number so when a Guthrie card barcode is scanned, it informs the lab staff that an extra hole punch (i.e. sample) is required for INGR1D. 3. Sample sent to **LGC lab** for genotyping. 4. A. The **neonatal screening lab** creates an <u>INGR1D ID</u> screening number and sends this to the **research portal** with the child’s date of birth and gender. B. The screening lab also exports to the **GPPAD database** using the <u>INGR1D ID,</u> the linked plate barcode and well ID. 5. The **research portal** sends to the **GPPAD database** the participant’s family history of T1D, the child’s date of birth and gender, and date of blood test, all linked to the <u>INGR1D ID</u> 6. **GPPAD database** generates a <u>GPPAD genetics ID</u> and sends this to **LGC**, linked to the Well ID and plate barcode. 7. **LGC** send the genotyping data back to the **GPPAD database** using the <u>GPPAD genetics ID</u> 8. **GPPAD database** uses the genotyping data from LGC with the family history, to work out the GRS and eligibility and inform the study team of the results linked to the <u>INGR1D ID</u>.

### Summary of Recruitment and Outcomes

From April 2018 to November 2020, 66% of women approached about INGR1D chose to participate, leading to a total of 15 660 babies being enrolled in the study. During this period 14 731 blood samples were processed; of whom 157 had confirmed positive results (>10% risk of multiple IA). Of these families 34 declined formal counselling about the positive result, and of the 124 families who undertook this counselling 49 agreed to take part in POInT. It is of note that 20 families were unable to participate in POInT due to lockdown restrictions. 107 (0.68%) of INGR1D’s 15 660 participants were withdrawn from the study. The most common reasons for withdrawal were a technical error with the sampling machine (36 participants), withdrawing consent or issues with consent (32 participants), and being out-of-area at the time the newborn blood spot was taken (27) (Figure 2).

**Figure 2.**
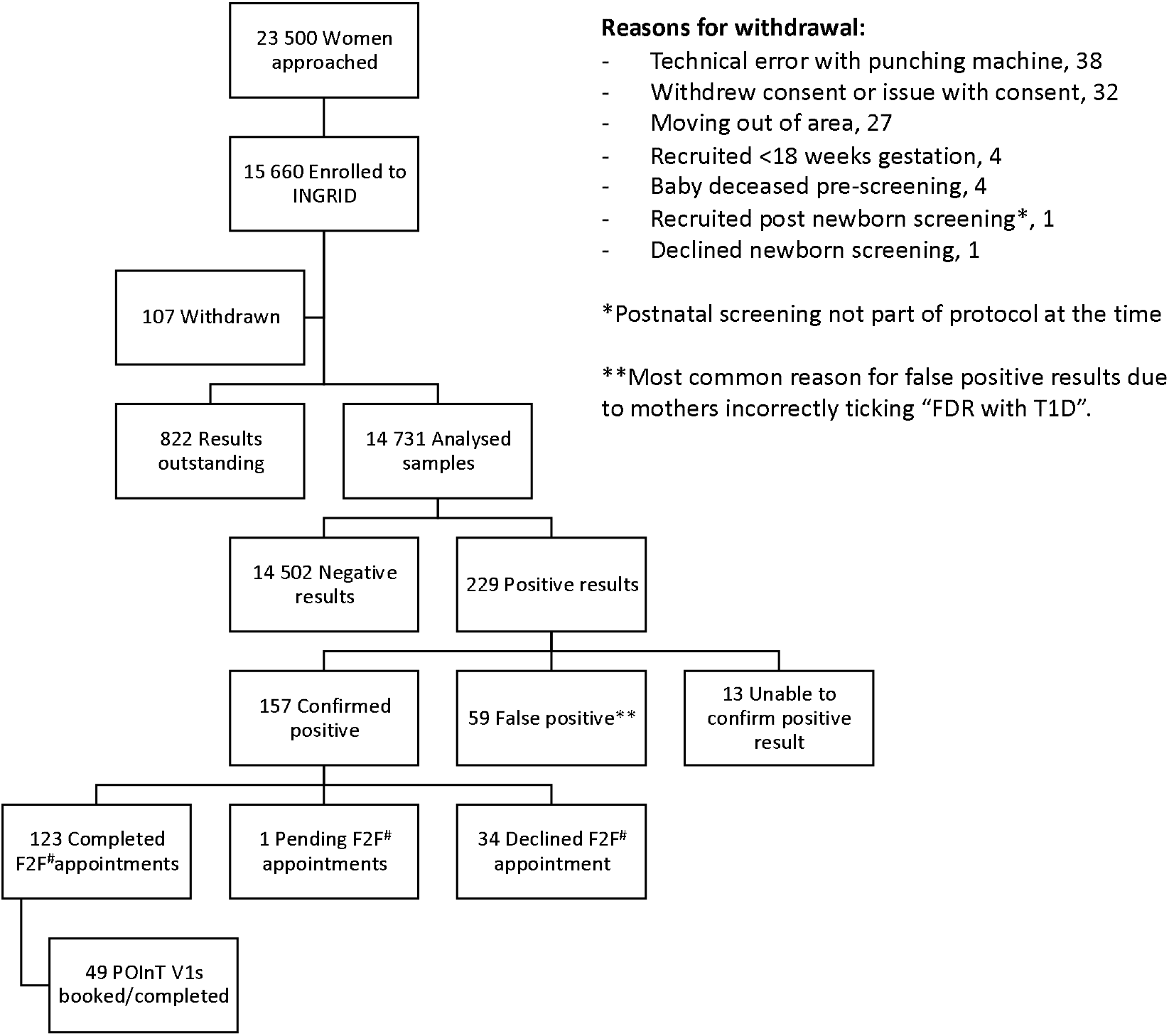
INGR1D and POInT accruals in first year (^#^ F2F - face-to-face)

Women reported that a principal reason for the successful recruitment to the screening study was the absence of any additional interventions. Of those women who declined screening, many had concerns regarding data protection. Some women feared their baby’s entire genome would be sequenced and its genetic data exploited by, for example, being sold to pharmaceutical companies. Others who declined did so based on the test’s accuracy; with a sensitivity of 25% some women worried about the value of a negative result. In addition, some stated that a predictive value of 10% meant that a high risk result could lead to unwarranted anxiety. Another barrier to women consenting to the study centred on understanding of disease risk. Many were falsely reassured by the fact they had no family history of T1D and therefore felt their baby would be low risk.

### GPPAD AND BEYOND

#### Future studies

Having developed and established this research platform, it is anticipated GPPAD will continue to utilise it for further T1D primary prevention studies. However, this methodology does not need to be solely restricted to T1D, genetic screening or interventions in the newborn. The model of antenatal recruitment (with the potential for supplementing the neonatal blood sample with maternal samples) also lends itself to exploring the impact of antenatal interventions, interrogation of the mother-foetus dyad and screening for at-risk population groups to offer postnatal primary interventions (e.g. to children born to mothers with gestational diabetes or pre-eclampsia, who have increased lifetime risks of diabetes, obesity and hypertension).

Furthermore, while INGR1D uses known SNPs to identify at-risk infants, the neonatal blood sample could be used to identify or utilise alternative biomarkers for a range of other diseases. Potential additional biomarkers include, but are not limited to, more expanded DNA analysis (exome, non-coding DNA, whole genome sequencing), epigenetic modifications (e.g. the methylome), transcriptomics, and non-genetic markers such as cytokines. If these data were collected at scale it would additionally be possible to establish “normal” reference ranges within this population group. Lastly, due to the timing of the day 5 blood spot, environmental influences and exposures to the child would be minimised and hence reduce further confounders.

#### Future Newborn Blood Spot Screening Programmes

In 2020, the UK celebrated 50 years since the inception of newborn blood spot screening for PKU. Over the past half a century however, the UK Newborn Screening Programme has evolved cautiously, taking three decades to include more than two screened diseases. Following a challenge by the chief medical officer in 2016, the UK NSC published a report in 2019 exploring the use of genomic medicine in all the screening programmes, including NBSSP.[29] The model used by GPPAD and described above, demonstrates that genomic technology can be integrated into a NBSSP and evaluated in parallel with routine testing without disrupting the screening pathway. These programmes have the potential to allow for early interventions prior to disease onset or progression. SMA represents a further example of a condition with a prognosis that can be dramatically improved through prompt identification and treatment.[30, 31] SMA can now be screened by polymerase chain reaction using a blood spot sample and already forms part of the screening programme in 18 states in the U.S. and in several regions in EU.[32-35]

Other such examples that perfectly match the screening criteria of Wilson and Jungner[36] include congenital myasthenia and Wilson’s disease. The experience garnered from GPPAD provides us with an opportunity to shift towards a much broader approach to newborn bloodspot screening which is in alignment with the UK’s intention to becoming a world leader in genomic medicine.

### GPPAD: a route towards 5 million genomes?

The UK declared its intention to become a pioneer in whole genome sequencing in 2012, launching the 100 000 genomes project and becoming the first healthcare system to launch a genomics medicine service.[37] This was established in recognition of the importance of genomic medicine as key to the future of personalised medicine. The project reached its goal in December 2018 and represents the world’s largest genome sequencing database with associated clinical data. The UK now wants to cement its position as world leader by sequencing a further 5 million genomes over the next 5 years in order to further advance our understanding of the link between genes and disease phenotypes.[38] The potential for incorporating whole genome sequencing for newborns would complement this approach by recruiting larger numbers of ‘healthy’ children. As described here, the successful introduction of INGR1D in the UK could serve as a template for the implementation of such a project by utilising the existing infrastructures of the NHS and the newborn screening programme, making population level genome-based research realisable.

Ultimately, the greatest asset from using surplus blood spots from newborn screening would be to undertake whole genome sequencing and have longitudinal life course follow-up using electronic patient health records. It would require very little input from the families and allow the generation of longitudinal disease phenotypes with rich data sources from primary and secondary care and non-health data. Instead of solely capturing cross-sectional data based on selective inclusion criteria, as adopted by the 100 000 genomes project, it would map the transition from disease onset to progression and treatment. Using a live electronic consent platform such as “Dynamic Consent” developed by Oxford University Innovation Technology (https://innovation.ox.ac.uk/licence-details/dynamic-consent/), participants could be approached about a study and give, review and change their consent preferences accordingly.

## Conclusion

We describe a methodology to recruit and identify newborns at increased risk of genetic disease by using antenatal consent and genetic analysis of surplus blood from the newborn blood spot screening programme. Over 66% of mothers approached agreed to take part, enabling enrolment of over 15500 babies in just over two and a half years. This demonstrates that not only is use of the NBSSP for genetic research both feasible and acceptable in a UK setting, but also that it does not interfere with the routine blood spot screening pathway. The INGR1D platform provides a model for future studies of this kind, with the potential to be expanded to ante-natal interventions and exploration of the mother-baby dyad, and represents the cutting edge of clinically relevant genetic research.

## Data Availability

Data mentioned in the manuscript is held either by the Oxford Vaccine Group, University of Oxford, or the Nuffield Department of Women's and Reproductive Health, University of Oxford

## Acknowledgements

The INGR1D study was financed as part of the GPPAD-02 study, which was funded by research grants from The Leona M. and Harry B. Helmsley Charitable Trust (Grant Numbers 2018PG-T1D022, 2018PG-T1D026).

Many thanks to the National Screening Programme Research Advisory Committee for granting approval for this study

With thanks also to the INGR1D study team: Patrick Bose, Wendy Byrne, Angelika Capp, Lotoyah Carty, Marie Cattle, Lianne Chapman, Edel Clare, Debbie Clarke, Chris Cleaver, Sarah Collins, Kate Dixon, Joy Edwards, Eleni Fotaki, Lisa Frankland, Mirella Garcia Corredera, Gemma Hawkins, Sue Johnston, Fidelma Lee, Joanna Mead, Jude Mossop, Sheila O′Connor, Dorota Pietrzak, Helen Price, Aparna Reddy, Suzanne Scanlon, Julie Tebbutt, Danielle Thornton, Sharon Westcar, Deborah Wilkinson, Catherine Young..

## Ethics

Approvals for this study were obtained from the National Screening Programme Research Advisory Committee, the Hampshire A Research Ethics Committee and the NHS Research and Development committees of the relevant NHS trusts.

## References

1. Downing, M. and R. Pollitt, Newborn bloodspot screening in the UK--past, present and future. Ann Clin Biochem, 2008. 45(Pt 1): p. 11–7.

2. England, P.H., Newborn Blood Spot Screening Programme in the UK. Data collection and performance analysis report 2016 to 2017. 2018.

3. IDF Diabetes Atlas. 2017, Internation Diabetes Federation.

4. Egro, F.M., Why is type 1 diabetes increasing? J Mol Endocrinol, 2013. 51(1):p. R1–13.

5. Patterson, C.C., et al., Incidence trends for childhood type 1 diabetes in Europe during 1989– 2003 and predicted new cases 2005–20: a multicentre prospective registration study. The Lancet, 2009. 373(9680):p. 2027–2033.

6. Mayer-Davis, E.J., et al., Incidence Trends of Type 1 and Type 2 Diabetes among Youths, 2002–2012. New England Journal of Medicine, 2017. 376(15):p. 1419–1429.

7. Group, D.E.R.I., Secular trends in incidence of childhood IDDM in 10 countries.. Diabetes, 1990. 39(7):p. 858–64.

8. Audit, N.P.D., National Paediatric Diabetes Audit 2016-17 Care Processes and Outcomes. 2018, Royal College of Paediatrics and Child Health.

9. Livingstone, S.J., et al., Estimated life expectancy in a Scottish cohort with type 1 diabetes, 2008-2010. JAMA, 2015. 313(1):p. 37–44.

10. Group, T.S., The Environmental Determinants of Diabetes in the Young (TEDDY) Study. Ann N Y Acad Sci, 2008. 1150:p. 1–13.

11. Rewers, M., et al., The Environmental Determinants of Diabetes in the Young (TEDDY) Study: 2018 Update. Curr Diab Rep, 2018. 18(12):p. 136.

12. Ziegler, A.G. and E. Bonifacio, Age-related islet autoantibody incidence in offspring of patients with type 1 diabetes. Diabetologia, 2012. 55(7):p. 1937–43.

13. Parikka, V., et al., Early seroconversion and rapidly increasing autoantibody concentrations predict prepubertal manifestation of type 1 diabetes in children at genetic risk. Diabetologia, 2012. 55(7):p. 1926–36.

14. Krischer, J.P., et al., The 6 year incidence of diabetes-associated autoantibodies in genetically at-risk children: the TEDDY study. Diabetologia, 2015. 58(5):p. 980–7.

15. Vafiadis, P., et al., Insulin expression in human thymus is modulated by INS VNTR alleles at the IDDM2 locus. Nat Genet, 1997. 15(3):p. 289–92.

16. Barratt, B.J., et al., Remapping the insulin gene/IDDM2 locus in type 1 diabetes. Diabetes, 2004. 53(7):p. 1884–9.

17. Du Toit, G., et al., Randomized trial of peanut consumption in infants at risk for peanut allergy. N Engl J Med, 2015. 372(9):p. 803–13.

18. Ziegler, A.G., et al., Primary prevention of beta-cell autoimmunity and type 1 diabetes - The Global Platform for the Prevention of Autoimmune Diabetes (GPPAD) perspectives. Mol Metab, 2016. 5(4):p. 255–262.

19. Ziegler, A.G., et al., Oral insulin therapy for primary prevention of type 1 diabetes in infants with high genetic risk: the GPPAD-POInT (global platform for the prevention of autoimmune diabetes primary oral insulin trial) study protocol. BMJ Open, 2019. 9(6):p. e028578.

20. Ahmed J. Delli, H.E.L., Sten-A. Ivarsson, Åke Lernmark Autoimmune type 1 diabetes in Textbook of Diabetes, C.S.C. RICHARD I.G. Holt, ALLAN Flyvbjerg, BARRY J. Goldstein, Editor. 2010, Wiley-Blackwell: Oxford.

21. Winkler, C., et al., Identification of infants with increased type 1 diabetes genetic risk for enrollment into Primary Prevention Trials—GPPAD-02 study design and first results. Pediatric Diabetes, 2019. 20(6):p. 720–727.

22. Bonifacio, E., Predicting type 1 diabetes using biomarkers. Diabetes Care, 2015. 38(6):p. 989–96.

23. Cooper, J.D., et al., Meta-analysis of genome-wide association study data identifies additional type 1 diabetes risk loci. Nat Genet, 2008. 40(12):p. 1399–401.

24. Lambert, A.P., et al., Absolute risk of childhood-onset type 1 diabetes defined by human leukocyte antigen class II genotype: a population-based study in the United Kingdom. J Clin Endocrinol Metab, 2004. 89(8):p. 4037–43.

25. Valdes, A.M., et al., Use of class I and class II HLA loci for predicting age at onset of type 1 diabetes in multiple populations. Diabetologia, 2012. 55(9):p. 2394–401.

26. Winkler, C., et al., Feature ranking of type 1 diabetes susceptibility genes improves prediction of type 1 diabetes. Diabetologia, 2014. 57(12):p. 2521–9.

27. Bonifacio, E., et al., Genetic scores to stratify risk of developing multiple islet autoantibodies and type 1 diabetes: A prospective study in children. PLoS Med, 2018. 15(4):p. e1002548.

28. Assfalg, R., et al., Oral insulin immunotherapy in children at risk for type 1 diabetes in a randomized trial. medRxiv, 2020:p. 2020.06.12.20129189.

29. Committee, U.N.S., Generation genome and the opportunities for screening programmes, P.H. England, Editor. 2019, PHE publications: London.

30. Dangouloff, T. and L. Servais, Clinical Evidence Supporting Early Treatment Of Patients With Spinal Muscular Atrophy: Current Perspectives. Ther Clin Risk Manag, 2019. 15:p. 1153–1161.

31. De Vivo, D.C., et al., Nusinersen initiated in infants during the presymptomatic stage of spinal muscular atrophy: Interim efficacy and safety results from the Phase 2 NURTURE study. Neuromuscul Disord, 2019. 29(11):p. 842–856.

32. Boemer, F., et al., Newborn screening for SMA in Southern Belgium. Neuromuscular disorders: NMD, 2019. 29(5):p. 343–349.

33. Dangouloff, T., et al., 244th ENMC international workshop: Newborn screening in spinal muscular atrophy May 10-12, 2019, Hoofdorp, The Netherlands. Neuromuscul Disord, 2020. 30(1):p. 93–103.

34. Vill, K., et al., One Year of Newborn Screening for SMA - Results of a German Pilot Project. J Neuromuscul Dis, 2019. 6(4):p. 503–515.

35. Kay, D.M., et al., Implementation of population-based newborn screening reveals low incidence of spinal muscular atrophy. Genet Med, 2020. 22(8):p. 1296–1302.

36. Petros, M., Revisiting the Wilson-Junger criteria: how can supplemental criteria guide public health in the era of genetic screening? Genet Med, 2012. 14(1): pp. 129-34

37. Investment, U.T.a., Genomics and personalised medicine: how partnership with the UK can transform healthcare, D.f.I. Trade, Editor. 2016.

38. Genome UK: the future of healthcare, D.o.H.a.S. Care, Editor. 2020.

